# Deep tissue sequencing improves genetic diagnostic yield in focal cortical dysplasia

**DOI:** 10.1101/2025.11.17.25340194

**Authors:** Breana Galea, Joshua Reid, Samuel Gooley, Tom Witkowski, Tara Lane, Sian Macdonald, Timothy E. Green, Zimeng Ye, Thiuni Adikari, Kristian Bulluss, Saul A. Mullen, Caitlin A. Bennett, Brialie Forster, Gabi Bradshaw, Wendi Lin, Wasanthi De Silva, Rosita B. Ramirez, Sattar Khoshkhoo, Sachin Gupta, Michael Krivanek, Kavitha Kothur, Deepak Gill, Kate Pope, Greta Gillies, Matthew Coleman, Wei-Shern Lee, Sarah M. Stephenson, Wirginia Maixner, A. Simon Harvey, Emma Macdonald-Laurs, Katherine B. Howell, Colleen D’Arcy, Paul J. Lockhart, Richard J. Leventer, Renata M. Kalnins, Jonathan Clark, Mark F. Bennett, Melanie Bahlo, Ingrid E. Scheffer, Piero Perucca, Samuel F. Berkovic, Michael S. Hildebrand

**Author notes:** Corresponding authors: Professor Michael S. Hildebrand: Epilepsy Research Centre, Level 1, Melbourne Brain Centre, 245 Burgundy Street, Heidelberg, Victoria 3084, Australia. Professor Samuel F. Berkovic: Epilepsy Research Centre, Level 1, Melbourne Brain Centre, 245 Burgundy Street, Heidelberg, Victoria 3084, Australia. These authors contributed equally to this work (Breana Galea, Joshua Reid, Samuel Gooley, Samuel F. Berkovic, Michael S. Hildebrand).

## Abstract

Focal cortical dysplasias (FCDs) are malformations of cortical development associated with drug-resistant focal epilepsy. We analysed surgical tissue from 28 consecutive cases recruited from adult and pediatric epilepsy surgery programs. We performed high-depth sequencing of lesional tissue, validated somatic variants using droplet digital PCR, and investigated genotype-phenotype correlations. A pathogenic or likely pathogenic variant was detected in 71% (n=20/28) of cases. Of these, six cases with FCDIIa or FCDIIb had germline variants in *NPRL3* (n=4) or *DEPDC5* (n=2). Somatic variants were identified in 50% (n=14/28) of cases. The genetic yield for FCDIIb was 85% of cases having a pathogenic mTOR pathway variant detected (n=12/14), and for FCDIIa 66% (n=6/9). This was achieved through high depth sequencing approaches that allowed detection of somatic variants with very low (down to 0.4%) variant allele fractions (VAFs). No pathogenic variants were detected in 3 cases with FCDI. 70% (n=18/26) of the cases with ≥12 months follow up experienced a favourable seizure outcome (Engel 1–2) following surgery. Of note, n=10 patients required repeat surgery to resect residual dysplasia. Determining a genetic diagnosis reveals aetiology and paves the way to precision therapies that may benefit those with FCD who do not respond to current treaments.

## 1. Introduction

Focal cortical dysplasia (FCD) is the most common histopathological diagnosis amongst children, and second most common amongst adults, with drug-resistant focal epilepsy requiring surgery (Blümcke et al., 2017). For these patients seizure freedom is often only achievable through surgical resection and in a subset a second surgery is required to remove residual dysplasia (Roessler et al., 2018). FCD comprises distinct subtypes with characteristic pathological and imaging features. FCDI is characterised by abnormal cortical lamination and lesions are often small and hard to detect radiologically. FCDIIa is characterised by cortical dyslamination with dysmorphic neurons, as is FCDIIb along with balloon cells; both typically show cortical thickening and grey–white blurring with or without a transmantle sign on MRI. FCDIII combines cortical dyslamination with an adjacent lesion, such as a glioneuronal tumour in FCDIIIb (Blümcke et al., 2011).

While genetic analyses of many pediatric surgical FCD cohorts have been described (ie., Baldassari et al., 2019; D’Gama et al., 2017), cohorts of predominantly adult cases have rarely been reported (Ferri et al., 2024). The major genetic etiologies of FCD are germline and somatic pathogenic variants of genes in the mechanistic (formerly mammalian) target of rapamycin (mTOR) pathway (Lim et al., 2014; Marsan & Baulac 2018; Baldassari et al., 2019; Najm et al., 2022; Macdonald-Laurs & Leventer et al., 2025). Reduced penetrance of some germline variants in these genes and case reports of biallelic variants supports a two-hit genetic model, similar to neoplasms, where inherited germline variants confer risk and interact with a second, somatic variant in brain tissue to initiate development of a dysplastic cortical malformation (Bennett et al., 2022; D’Gama et al., 2017; Lee et al., 2019; Ribierre et al., 2018).

Across surgical cohorts, FCD subtype is one of the strongest predictors of postoperative seizure outcome. Consistently, patients with FCDII, and particularly FCDIIb, have better postoperative seizure outcomes than FCDI. Post-surgery seizure outcomes were not correlated with genetic diagnosis in these cohorts (Tassi et al., 2002; Krsek et al., 2009, Lamberink et al., 2020).

Here, we aimed to (i) improve the genetic diagnostic yield for patients with FCD and drug-resistant focal epilepsy requiring surgery by interrogating deep tissue sequencing data for somatic variants, and (ii) determine the impact of a genetic diagnosis on seizure outcomes post-surgery.

## 2. Materials & Methods

### Cohort and Sample Collection

Our cohort comprised 28 cases with drug-resistant focal epilepsy where histopathology of surgically resected brain tissue confirmed FCD (see **Supplementary Table 1** for patient list). Cases were recruited from three epilepsy surgery programs. Written informed consent was obtained for all cases with approval from Human Research Ethics Committees at Austin Health, Melbourne, The Royal Children’s Hospital, Melbourne, The Westmead Children’s Hospital, Sydney and The Walter and Eliza Hall Institute of Medical Research, Melbourne.

Surgically resected fresh-frozen (FF) or formalin-fixed paraffin-embedded (FFPE) brain tissue was collected from all cases. For further analysis of variants, whole blood and buccal samples and skin biopsies were also collected where possible. Genomic DNA was extracted using commercially available kits, including the QIAamp DNA Blood Maxi Kit for whole blood and DNA Genotek prepIT.L2P for buccal samples. The QIAamp DNA FFPE Tissue Kit was used to extract all fresh-frozen brain specimens and one archival brain FFPE specimen including the deparaffinisation step.

### High-Depth Exome Sequencing

DNA libraries were prepared using the SureSelect Human All Exon V6 Kit (Agilent) and sequenced on either an Illumina NovaSeq 6000 or X Plus system at BGI Genomics (Hong Kong) with 150bp paired-end reads, targeting 200–400x exome-wide coverage per sample. Read alignment to the GRCh38 human reference genome was performed using BWA-MEM, followed by realignment and recalibration with GATK. Variant calling was performed using the Sarek pipeline from Nextflow Core (Garcia, Juhos & Larsson et al., 2020), which implements multiple somatic callers; Mutect2, Strelka2 and FreeBayes.

### High-Depth Panel Sequencing

DNA libraries were prepared using the SureSelect XT HS2 DNA Kit (Agilent) with duplex unique molecular identifiers (UMIs) and a custom gene panel targeting 140 genes, including 36 Ras/Raf/MAPK pathway genes, 12 mTOR pathway genes, 7 other focal epilepsy genes and 85 additional cancer genes (see **Supplementary Table 2** for gene list). Sequencing was performed on an Illumina NextSeq 1000 platform at Austin Pathology with 150bp paired-end reads. Read alignment to the GRCh38 human reference genome and variant calling were performed using the DRAGEN DNA Pipeline on Illumina Basespace (v4.3.13). UMI tags were used for read collapsing during alignment, achieving a median consensus depth of 582x per sample across panel positions.

### Variant Prioritisation

Exome variant analysis was restricted to genes in a curated panel derived from: Genes4Epilepsy (v2025-03, Oliver et al., 2023), KEGG Ras and mTOR signaling pathways (Kanehisa et al., 2025; Kanehisa, 2019; Kanehisa & Goto, 2000), PanelApp Australia Malformations of Cortical Development Superpanel (v4.65, Martin & Williams et al., 2019), and the Austin Pathology panel above. Exome variants were required to have ≥2 alternate reads on each strand (forward and reverse). Gene panel variants were filtered to target regions and required at least 2 supporting consensus reads. Quality filtered variants were annotated with Ensembl Variant Effect Predictor (VEP), CADD pathogenicity scores, gnomAD population allele frequencies, and ClinVar database entries. Pathogenicity was evaluated using Franklin by Genoox (Qiagen, Hilden, Germany) with pathogenic and likely pathogenic variants prioritised for validation. Candidate variants were visualised in Integrative Genomics Viewer (IGV) for manual quality assessment.

### Droplet Digital PCR

Low frequency somatic variants were validated utilising Bio-Rad droplet digital PCR (ddPCR) with TaqMan® SNP Genotyping Assays customed ordered from Thermo Fisher Scientific (Waltham, MA). Fluorescently-labelled probes were added to ddPCR Supermix for probes (no dUTPs) (Bio-Rad, Hercules, CA) and mixed with 10ng of DNA derived from surgically resected fresh-frozen brain tissue. Generation of droplets, PCR cycling and droplet reading were performed according to the manufacturer’s instructions. Graphical outputs were created using RStudio.

### Sanger Sequencing

Germline and high frequency somatic variants were validated using PCR and Sanger sequencing. Gene variants were amplified using gene-specific primers (oligonucleotide sequences available on request) designed to the reference human gene transcripts (NCBI Gene). Amplification reactions were cycled using a standard protocol on a Veriti Thermal Cycler (Applied Biosystems, Carlsbad, CA). Bidirectional sequencing was completed with a BigDye^TM^ v3.1 Terminator Cycle Sequencing Kit (Applied Biosystems), according to the manufacturer’s instructions. Sequencing products were resolved using a 3730xl DNA Analyzer (Applied Biosystems). All sequencing chromatograms were compared to the published cDNA sequence; nucleotide changes were detected using Codon Code Aligner (CodonCode Corporation, Dedham, MA).

### Immunostaining

Archival FFPE tissue sections from case 12 with a somatic *MTOR* variant were immunostained according to a published protocol (Baybis et al., 2004). Blocking buffer consisted of 2% fetal bovine serum (FBS) in 1X phosphate-buffered saline (PBS). FFPE tissue slides were immersed in citrate-based antigen unmasking solution (#H-3300, Vector Laboratories, Newark, CA) and incubated overnight with Phospho-S6 Ribosomal Protein (Ser235/236) (D57.2.2E) XP® Rabbit mAb (1:1000) (#4858, Cell Signalling Technology, Danvers, MA) at 4°C. The slides were then incubated in secondary antibody Goat Anti-Rabbit IgG H&L (HRP) (1:1000) (#ab6721, Abcam, Cambridge, UK) for 1 hour at room temperature. Immunoreactivity was visualised utilising the VECTASTAIN® ABC-HRP Kit, Peroxidase (Standard) (#PK-4000, Vector Laboratories) and DAB Substrate Kit, Peroxidase (HRP), with Nickel, (3,3’-diaminobenzidine) (#SK-4100, Vector Laboratories), counterstaining with haematoxylin for 10 seconds from the Hematoxylin and Eosin Stain Kit (#H-3502, Vector Laboratories). Commercial kits were used according to the manufacturer’s recommendations.

### Reverse transcription polymerase chain reaction (RT-PCR)

RNA was extracted from brain tissue or cultured fibroblasts using the Qiagen AllPrep DNA/RNA Kit. cDNA synthesis was performed with the SuperScript™ III First-Strand Synthesis System using random hexamer primers. Region-specific primers (Supplementary Table 3) were used to amplify cDNA by PCR. RT-PCR products were analyzed by gel electrophoresis and Sanger sequencing to assess variant impact on RNA splicing.

### Phenotyping and statistical analysis

Seizure outcome was classified using the Engel scale (Engel et al., 1993). A single Engel class at last follow-up was assigned based on the postoperative seizure course from the most recent epilepsy surgery. An Engel class was only assigned if the patient had ≥12 months follow up from the date of their most recent resective surgery. A favourable seizure outcome was defined as Engel class 1 or 2 and an unfavourable seizure outcome was defined as Engel class 3 or 4.

In the phenotyping and genotype–phenotype correlation analyses, descriptive statistics were used to summarize clinical characteristics including age of seizure onset and age at first surgery. Comparisons between groups (e.g., germline vs. somatic variants, presence vs. absence of a pathogenic variant) were evaluated using Fisher’s exact test for categorical outcomes such as surgical response, due to small sample sizes. For continuous variables like age of seizure onset, group comparisons were performed using the Mann–Whitney U test. Statistical significance was defined using a p-value threshold of <0.05.

## 3. Results

### Clinical Phenotypes

All patients (n=28) underwent resective surgery for treatment of drug-resistant focal epilepsy. Focal cortical dysplasia was confirmed histopathologically in each case with the most common subtype being FCDIIb (n=14 patients), followed by FCDIIa (n=9), FCDI (n=3) and FCDIIIb (n=2). The FCDs were most frequently located in the frontal lobe (n=15), followed by the parietal lobe (n=6), temporal lobe (n=5), occipital lobe (n=1) and a hemispheric dysplasia (n=1); the latter case has previously been reported (Leventer., et al 2015). Of note, the FCD involved either the frontal or parietal cortex in 87% (n=20/23) cases with FCDII (a or b). Across the cohort, the median age of seizure onset was 5.5 years (range: 6 weeks to 30 years), and the median age of first epilepsy surgery was 25 years (6 months to 63 years).

At least 12 months of post-operative follow up surgical outcome data (from the date of the most recent surgery) was available for n=26 cases with a mean of 6.2 years of follow up (1–11years). 70% (n=18/26) experienced a favourable seizure outcome (Engel 1–2). Ten patients underwent repeat surgery to resect residual dysplasia. Of these ten cases requiring multiple surgeries, five had a favourable seizure outcome after the repeat surgery, four had an unfavourable outcome and one had < 12 months follow up since the repeat surgery. Patients with FCDIIb tended to have better surgical outcomes than those with FCDI or FCDIIa. Favourable seizure outcomes were seen in 83% of FCD IIb cases (n=10/12), compared with 33% of FCDI cases (n=1/3) and 44% of FCDIIa cases (n=4/9). When FCDIIb was compared with all other FCD subtypes combined, this difference showed a non-significant trend (p = 0.089). However, these comparisons are underpowered due to small subgroup sizes. The n=2 patients with FCDIIIb both had a favourable seizure outcome from surgery.

### Genetic Findings

At least one pathogenic variant was detected in 71% (n=20/28) of cases. Germline pathogenic variants were identified in 21% (n=6/28) of cases, comprising *NPRL3* (n=4) and *DEPDC5* (n=2). The *NPRL3* splice site variant (case 2) was shown to cause skipping of exon 5 in brain-derived RNA (see **Supplementary Figure 1**). The germline *DEPDC5* variant in case 6 was part of a single nuclei RNA sequencing study of FCD that showed disruption of glutamate and GABA-A signalling pathways in variant-carrying neurons (Bizzotto., et al 2025). Somatic variants were found in 50% (n=14/28) of cases, consisting of *MTOR* (n=12) and *BRAF* (n=2). Of the n=12 cases with a pathogenic somatic *MTOR* variant, the median variant allele fraction (VAF) was 2.4% with a range of 0.4–8.0%. In one FCDIIb case with a pathogenic somatic *MTOR* variant at a VAF of 2.4% (case 12), we were able to confirm hyperactivation of the mTOR pathway with immunostaining of lesional tissue demonstrating increased cytoplasmic phosphorylation of ribosomal S6 protein (p-S6) in dysmorphic neurons **(Figure 1)**. Somatic variants were absent in paired blood or buccal samples in all cases with samples available. Two (n=2/28) double-hit germline-somatic cases were identified, one with a pathogenic somatic *MTOR* variant and a candidate germline variant in *EIF4ENIF1* (case 18), and one with a pathogenic germline *NPRL3* and a candidate somatic variant in *WNT2* (case 4). Case 3 is sibling of case 4 and also carries the germline *NPRL3* variant. We have previously reported this finding in cases 3 and 4 (Bennett et al., 2022). **(Table 1)**.

**Figure 1.**
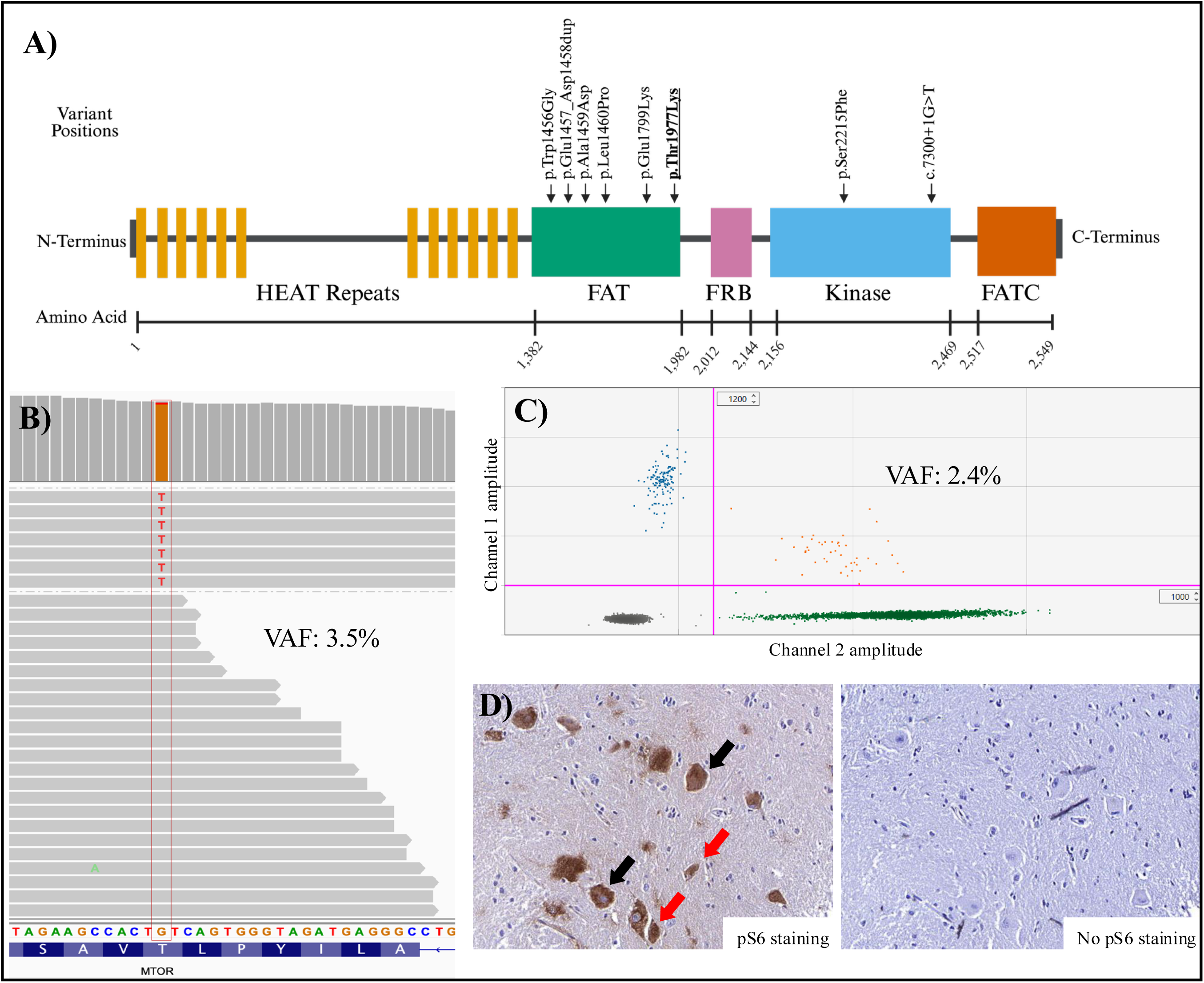
Molecular and functional results for low frequency MTOR variant (c.5930C>A, p.Thr1977Lys) found in genomic DNA of case 12 with FCD IIb. VAF: variant allele fraction. p-S6: 25hosphor-S6. Top-left: Diagram of MTOR variant. Created using BioRender. Top-right: Screenshot of exome data results in fresh-frozen brain tissue using Integrative Genomics Viewer. The MTOR variant is present at a VAF of 3.5% (7 reads in a total of 193 reads). The box indicates the position of the nucleotide change. Middle-left: Graph representing droplet digital PCR (ddPCR) validation in fresh-frozen brain tissue at a VAF of 2.4%, with the y-axis corresponding to channel 1 and the x-axis corresponding to channel 2. Results represent mutant alleles (blue droplets), wildtype alleles (green droplets), wildtype and mutant alleles (orange droplets) and no DNA template (grey droplets). Bottom: Two histology images showing FFPE tissue sections with or without p-S6 staining. Red arrows show dysmorphic neurons and black arrows show balloon cells. Images cropped from whole slide scans taken at 40X magnification on the PALM Laser Dissector System Axiovert 200M (ZEISS, Oberkochen, Germany).

**Table 1.**
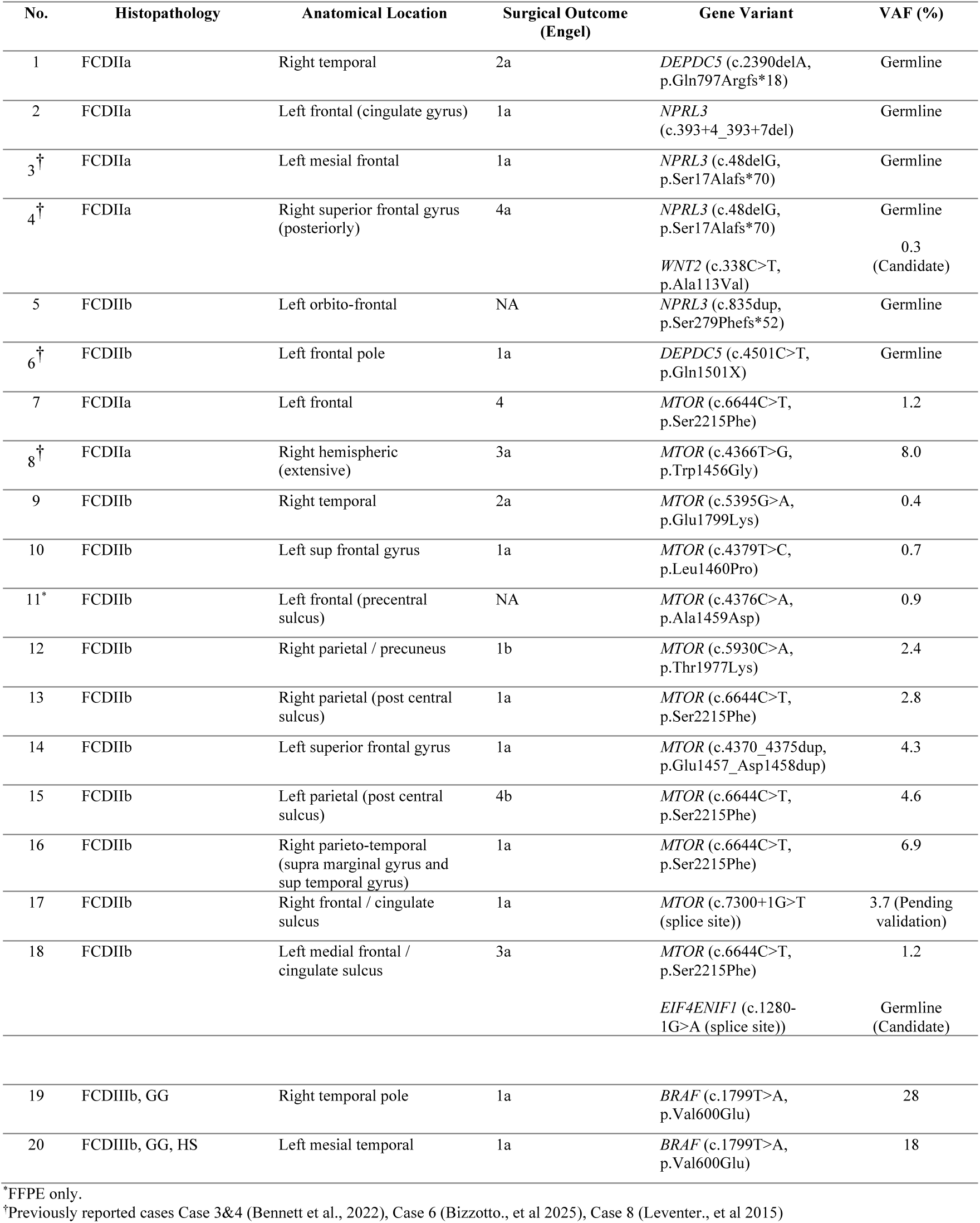
Summary of findings in novel and established epilepsy genes. VAF: variant allele fraction. FCD: focal cortical dysplasia. VUS: variant of unknown significance. TSC: tuberous sclerosis. GG: ganglioglioma. HS: hippocampal sclerosis. NA: Not applicable (<12 months follow up from most recent surgery)

| No. | Histopathology | Anatomical Location | Surgical Outcome (Engel) | Gene Variant | VAF (%) |
| --- | --- | --- | --- | --- | --- |
| 1 | FCDIIa | Right temporal | 2a | *DEPDC5* (c.2390delA, p.Gln797Argfs*18) | Germline |
| 2 | FCDIIa | Left frontal (cingulate gyrus) | 1a | *NPRL3* (c.393+4_393+7del) | Germline |
| 3<sup>†</sup> | FCDIIa | Left mesial frontal | 1a | *NPRL3* (c.48delG, p.Ser17Alafs*70) | Germline |
| 4<sup>†</sup> | FCDIIa | Right superior frontal gyrus (posteriorly) | 4a | *NPRL3* (c.48delG, p.Ser17Alafs*70)<br>*WNT2* (c.338C>T, p.Ala113Val) | Germline<br>0.3 (Candidate) |
| 5 | FCDIIb | Left orbito-frontal | NA | *NPRL3* (c.835dup, p.Ser279Phefs*52) | Germline |
| 6<sup>†</sup> | FCDIIb | Left frontal pole | 1a | *DEPDC5* (c.4501C>T, p.Gln1501X) | Germline |
| 7 | FCDIIa | Left frontal | 4 | *MTOR* (c.6644C>T, p.Ser2215Phe) | 1.2 |
| 8<sup>†</sup> | FCDIIa | Right hemispheric (extensive) | 3a | *MTOR* (c.4366T>G, p.Trp1456Gly) | 8.0 |
| 9 | FCDIIb | Right temporal | 2a | *MTOR* (c.5395G>A, p.Glu1799Lys) | 0.4 |
| 10 | FCDIIb | Left sup frontal gyrus | 1a | *MTOR* (c.4379T>C, p.Leu1460Pro) | 0.7 |
| 11* | FCDIIb | Left frontal (precentral sulcus) | NA | *MTOR* (c.4376C>A, p.Ala1459Asp) | 0.9 |
| 12 | FCDIIb | Right parietal / precuneus | 1b | *MTOR* (c.5930C>A, p.Thr1977Lys) | 2.4 |
| 13 | FCDIIb | Right parietal (post central sulcus) | 1a | *MTOR* (c.6644C>T, p.Ser2215Phe) | 2.8 |
| 14 | FCDIIb | Left superior frontal gyrus | 1a | *MTOR* (c.4370_4375dup, p.Glu1457_Asp1458dup) | 4.3 |
| 15 | FCDIIb | Left parietal (post central sulcus) | 4b | *MTOR* (c.6644C>T, p.Ser2215Phe) | 4.6 |
| 16 | FCDIIb | Right parieto-temporal (supra marginal gyrus and sup temporal gyrus) | 1a | *MTOR* (c.6644C>T, p.Ser2215Phe) | 6.9 |
| 17 | FCDIIb | Right frontal / cingulate sulcus | 1a | *MTOR* (c.7300+1G>T (splice site)) | 3.7 (Pending validation) |
| 18 | FCDIIb | Left medial frontal / cingulate sulcus | 3a | *MTOR* (c.6644C>T, p.Ser2215Phe)<br>*EIF4ENIF1* (c.1280-1G>A (splice site)) | 1.2<br>Germline (Candidate) |
| 19 | FCDIIIb, GG | Right temporal pole | 1a | *BRAF* (c.1799T>A, p.Val600Glu) | 28 |
| 20 | FCDIIIb, GG, HS | Left mesial temporal | 1a | *BRAF* (c.1799T>A, p.Val600Glu) | 18 |

\*FFPE only.
<sup>†</sup>Previously reported cases Case 3&4 (Bennett et al., 2022), Case 6 (Bizzotto., et al 2025), Case 8 (Leventer., et al 2015)

### Genotype–Phenotype Correlations

Pathogenic or likely pathogenic mTOR pathway variants were identified in 85% (n=12/14) of cases with FCDIIb (2 germline, 10 somatic) and in 66% (n=6/9) cases with FCDIIa (4 germline, 2 somatic). There was marked genetic homogeneity in the FCDIIb cases with a pathogenic somatic variant in the *MTOR* gene being detected in 71% (n=10/14) of cases, compared to 22% (n=2/9) of FCDIIa, and 0% (n=0/7) of other dysplasia types (FCDI, FCDIII) (p=0.01). No pathogenic variants were identified in the three cases with FCDI. Both cases with FCDIIIb had a ganglioglioma with the highly recurrent *BRAF* V600E variant.

Of the n=6 cases with a pathogenic or likely pathogenic germline mTOR pathway variant, the median age of seizure onset was 1.3 years (range 0.3–21 years). This was earlier than the median age of 6 years (range 0.1–13 years) in the n=12 cases with a somatic mTOR pathway variant. However, this difference was not statistically significant (p=0.38). Patients with a germline mTOR pathway variant tended to undergo surgery at a younger age (median of 6.5 years; range 2–49 years) compared to a median of 25 years (range 0.5–63 years) in those with a somatic variant. This difference was also not statistically significant (p=0.31) and both of these comparisons are limited by small subgroup sizes.

Patients with a pathogenic variant (germline or somatic) detected tended to have a better seizure outcome after surgery compared to those without. Of those with a pathogenic variant, 72% (n=13/18) had a favourable seizure outcome, compared to 50% (n=4/4) with no variant detected. This difference was not statistically significant (p=0.32) and the analysis is underpowered due to small subgroup sizes. Of the ten patients who underwent repeat surgeries, five had a pathogenic variant and five did not.

We were underpowered to explore whether variant allele fractions (VAFs) affected surgical outcomes in the n=11 patients with a somatic *MTOR* variant and surgical follow up data available. However, similar VAFs were seen in those with favourable and unfavourable outcomes after surgery with VAFs ranging from 0.4–6.9% in those with a favourable surgical outcome and 1.2–8% in those with an unfavourable outcome. Median VAFs were 2.8% and 2.9%, respectively, with no significant difference between groups (p=0.57).

## 4. Discussion

Our high-depth sequencing approach optimised for detection of somatic variants at very low variant allele fractions resulted in a high yield of genetic diagnosis in this cohort, particularily for FCDIIb with a yield of 85%. We identified five pathogenic somatic *MTOR* variants below 1.5% VAF, with one case as low as 0.4% (case 9). All of these low VAF variants were independently validated by ddPCR. This capacity to resolve low-level mosaicism underlied our higher yield relative to earlier FCDII series that reported somatic mTOR pathway variants in 35–60% of cases with median VAFs ∼3–5% (Baldassari et al., 2019; Lai et al., 2022). More recent studies have shown that very-low VAF variants are an important cause of FCDII, as we confirm. For example, enrichment of p-S6–positive lesional cells followed by deep sequencing revealed mTOR pathway variants with VAFs <0.2% (Kim et al., 2023) and ultra-deep sequencing has since detected mosaicism down to 0.07% in bulk brain tissue from FCDII patients (Kim et al., 2024). Collectively, these data suggest that genetic testing for FCD should employ sequencing strategies that reliably detect variants with allele fractions well below 1%.

Double-hit germline-somatic variants in FCD are rarely reported and typically occur in the same gene. (Lee et., 2019; Ribierre et al., 2018). Noteably, we detected dual-pathway, double-hit germline-somatic variants in 2 cases (cases 4 and 18). Both cases had a single mTOR pathway gene variant (germline *NPRL3* or somatic *MTOR*) with a candidate variant from another pathway (somatic *WNT2* or germline *EIFENIF4*).

FCDIIb was the most common histopathological subtype, followed by FCDIIa, with the majority (87%, n=20/23) of FCDII (a or b) being located in the frontal or parietal lobes, consistent with findings from other FCD cohorts (Macdonald-Laurs et al., 2024). Those with a pathogenic germline mTOR pathway variant tended to have an earlier age of seizure onset compared to those with a somatic mTOR pathway variant, which is a phenomenon recently reported in patients with bottom-of-sulcus dysplasias (Macdonald-Laurs et al., 2024), although we were underpowered to confirm this difference was significant in our cohort. Patients with FCDIIb tended to have better surgical outcomes than those with FCDI or FCDIIa. We were also underpowered to show that this was a statistically significant difference in our cohort, but this phenomenon has been reported in larger cohorts (Tassi et al., 2002; Krsek et al., 2009, Lamberink et al., 2020).

In summary, the frequent detection of mTOR pathway gene variants in this study aligns with recent work in this field that has confirmed mTOR pathway dysregulation as the dominant cause of FCD. Recent advances in ultra-deep sequencing is enabling accurate detection of somatic variants at ultra-low VAFs and cutting-edge single-cell genotyping and transcriptomic techniques are rapidly expanding our understanding of the developmental timing of these variants, the specific cell types involved and how these affected cells are interacting with surrounding brain tissue to produce seizures (Kim et al., 2024; Bizzotto et al., 2025). This integration of genetic diagnosis with mechanistic biology is paving the way for the emerging use of FDA-approved mTOR inhibitors like everolimus for individuals who are positive for an mTOR pathway gene variant and who continue to have seizures despite current treatments.

## Author statement

The manuscript has been read and approved by all authors. Artificial Intelligence was not used in any stage of the generation or editing of this article. All funding sources for this work have been reported in the declaration statement.

## CRediT authorship contribution statement

**Breana Galea:** Data Curation, Formal Analysis, Validation, Writing – Original Draft.

**Joshua Reid:** Data Curation, Formal Analysis.

**Samuel Gooley:** Data Curation, Formal Analysis, Validation, Writing – Original Draft.

**Tom Witkowski:** Data Curation, Formal Analysis, Validation.

**Tara Lane:** Data Validation.

**Sian Macdonald:** Data Curation,Validation.

**Timothy Green:** Data Curation, Formal Analysis, Validation.

**Zimeng Ye:** Validation.

**Thiuni Adikari**: Data Curation, Formal Analysis, Validation.

**Kristian Bulluss:** Data Curation.

**Saul Mullen:** Data Curation.

**Caitlin Bennett:** Data Curation.

**Brialie Forster:** Data Curation.

**Gabi Bradshaw:** Data Curation, Formal Analysis.

**Wendi Lin:** Data Curation, Formal Analysis.

**Wasanthi De Silva:** Data Curation, Formal Analysis.

**Rosita Ramirez**: Data Curation, Formal Analysis.

**Sattar Khoshkhoo**: Data Curation, Formal Analysis, Funding Acquisition.

**Sachin Gupta:** Data Curation.

**Michael Krivanek:** Data Curation.

**Kavitha Kothur:** Data Curation.

**Deepak Gill:** Data Curation.

**Kate Pope:** Data Curation.

**Greta Gillies:** Data Curation.

**Matthew Coleman:** Data Curation, Formal Analysis, Validation.

**Wei-Shern Lee**: Data Curation, Formal Analysis, Validation.

**Sarah Stephenson:** Data Curation, Formal Analysis, Validation.

**Wirginia Maixner:** Data Curation.

A. **Simon Harvey:** Data Curation.

**Emma Macdonald-Laurs:** Data Curation, Formal Analysis.

**Katherine Howell:** Data Curation, Formal Analysis.

**Colleen D’Arcy:** Data Curation, Formal Analysis.

**Paul Lockhart:** Data Curation, Formal Analysis, Funding Acquisition.

**Richard Leventer:** Data Curation, Formal Analysis, Funding Acquisition.

**Renata Kalnins:** Data Curation, Formal Analysis.

**Jonathan Clark:** Data Curation, Formal Analysis, Funding Acquisition.

**Mark F. Bennett:** Data Curation, Formal Analysis.

**Melanie Bahlo:** Data Curation, Formal Analysis, Funding Acquisition.

**Ingrid E. Scheffer:** Data Curation, Formal Analysis, Funding Acquisition.

**Piero Perucca:** Data Curation, Formal Analysis, Funding Acquisition.

**Samuel F. Berkovic:** Conceptualization, Data Curation, Formal Analysis, Funding Acquisition, Project Administration, Supervision, Validation, Writing – Original Draft, Writing – Review and Editing.

**Michael S. Hildebrand:** Conceptualization, Data Curation, Formal Analysis, Funding Acquisition, Project Administration, Supervision, Validation, Writing – Original Draft, Writing – Review and Editing.

## Data availability

Gene variants have been deposited in the publicly accessible Leiden Open Variation Database v3.0 (https://www.lovd.nl/), with a direct link provided upon request.

## Acknowledgements

We thank all patients for their participation in the study. The authors gratefully acknowledge the Biological Optical Microscopy Platform for their support and assistance in this work.

