## Supplementary Material for "Deep tissue sequencing improves genetic diagnostic yield in focal cortical dysplasia"

**Supplementary Table 1.** Summary of findings in novel and established epilepsy genes, including unsolved cases. ACMG: American College of Medical Genetics and Genomics. gnomAD: Genome Aggregation Database. VAF: variant allele fraction. FCD: focal cortical dysplasia. VUS: variant of unknown significance. TSC: tuberous sclerosis. GG: ganglioglioma. HS: hippocampal sclerosis. NA: Not applicable (<12months follow up from most recent surgery)

| No. | Hospital | Histopathology | Anatomical Location | Surgical Outcome (Engel) | Gene Variant | ACMG | gnomAD (%) | VAF (%) | NM Code | ENST Code | Tissue Sequencing Method | Solved |
|---|---|---|---|---|---|---|---|---|---|---|---|---|
| 1 | RCH | FCDIIa | Right temporal | 2a | *DEPDC5* (c.2390delA, p.Gln797Argfs*18) | Likely pathogenic | 0 | Germline | NM_001242896.3 | ENST00000400246.7 | Exome | Y |
| 2 | RCH | FCDIIa | Left frontal (cingulate gyrus) | 1a | *NPRL3* (c.393+4_393+7del) | Likely pathogenic | 0.001 | Germline | NM_001077350.3 | ENST00000611875.5 | Exome | Y |
| 3† | Westmead | FCDIIa | Left mesial frontal | 1a | *NPRL3* (c.48delG, p.Ser17Alafs*70) | Likely pathogenic | 0 | Germline | NM_001077350.3 | ENST00000611875.5 | Exome | Y |
| 4† | Westmead | FCDIIa | Right superior frontal gyrus (posteriorly) | 4a | *NPRL3* (c.48delG, p.Ser17Alafs*70) | Likely pathogenic | 0 | Germline | NM_001077350.3 | ENST00000611875.5 | Exome | Y |
| | | | | | *WNT2* (c.338C>T, p.Ala113Val) | VUS | 0 | 0.3 (Candidate) | NM_003391.3 | ENST00000265441.8 | | |
| 5 | Austin | FCDIIb | Left orbito-frontal | NA | *NPRL3* (c.835dup, p.Ser279Phefs*52) | Pathogenic | 0 | Germline | NM_001077350.3 | ENST00000611875.5 | Exome | Y |
| 6† | Austin | FCDIIb | Left frontal pole | 1a | *DEPDC5* (c.4501C>T, p.Gln1501X) | Pathogenic | 0 | Germline | NM_001242896.3 | - | Exome | Y |
| 7 | Westmead | FCDIIa | Left frontal | 4 | *MTOR* (c.6644C>T, p.Ser2215Phe) | Pathogenic | 0 | 1.2 | NM_004958.4 | ENST00000361445.9 | Exome, Gene panel | Y |
| 8† | RCH | FCDIIa | Right hemispheric (extensive) | 3a | *MTOR* (c.4366T>G, p.Trp1456Gly) | Likely pathogenic | 0 | 8.0 | NM_004958.4 | ENST00000361445.9 | Exome | Y |
| 9 | Austin | FCDIIb | Right temporal | 2a | *MTOR* (c.5395G>A, p.Glu1799Lys) | Pathogenic | 0 | 0.4 | NM_004958.4 | ENST00000361445.9 | Exome | Y |
| 10 | Austin | FCDIIb | Left sup frontal gyrus | 1a | *MTOR* (c.4379T>C, p.Leu1460Pro) | Pathogenic | 0 | 0.7 | NM_004958.4 | ENST00000361445.9 | Exome, Gene panel | Y |
| 11* | Austin | FCDIIb | Left frontal (precentral sulcus) | NA | *MTOR* (c.4376C>A, p.Ala1459Asp) | Pathogenic | 0 | 0.9 | NM_004958.4 | ENST00000361445.9 | Exome, Panel | Y |
| 12 | Austin | FCDIIb | Right parietal / precuneus | 1b | *MTOR* (c.5930C>A, p.Thr1977Lys) | Pathogenic | 0 | 2.4 | NM_004958.4 | ENST00000361445.9 | Exome | Y |
| 13 | Austin | FCDIIb | Right parietal (post central sulcus) | 1a | *MTOR* (c.6644C>T, p.Ser2215Phe) | Pathogenic | 0 | 2.8 | NM_004958.4 | ENST00000361445.9 | Exome | Y |
| 14 | Austin | FCDIIb | Left superior frontal gyrus | 1a | *MTOR* (c.4370_4375dup, p.Glu1457_Asp1458dup) | Likely pathogenic | 0 | 4.3 | NM_004958.4 | ENST00000361445.9 | Exome, Gene panel | Y |

| | | | | | | | | | | | |
|---|---|---|---|---|---|---|---|---|---|---|---|
| 15 | Austin | FCDIIb | Left parietal (post central sulcus) | 4b | *MTOR* (c.6644C>T, p.Ser2215Phe) | Pathogenic | 0 | 4.6 | NM_004958.4 | ENST00000361445.9 | Exome | Y |
| 16 | Austin | FCDIIb | Right parieto-temporal (supra marginal gyrus and sup temporal gyrus) | 1a | *MTOR* (c.6644C>T, p.Ser2215Phe) | Pathogenic | 0 | 6.9 | NM_004958.4 | ENST00000361445.9 | Exome | Y |
| 17 | Austin | FCDIIb | Right frontal / cingulate sulcus | 1a | *MTOR* (c.7300+1G>T (splice site)) | VUS | 0 | 3.7 (Pending validation) | NM_004958.4 | ENST00000361445.9 | Exome | Y |
| 18 | Austin | FCDIIb | Left medial frontal / cingulate sulcus | 3a | *MTOR* (c.6644C>T, p.Ser2215Phe) | Pathogenic | 0 | 1.2 | NM_004958.4 | ENST00000361445.9 | Exome | Y |
| | | | | | *EIF4ENIF1* (c.1280-1G>A (splice site)) | VUS | 0 | Germline (Candidate) | NM_019843.4 | ENST00000330125.10 | | |
| 19 | Austin | FCDIIIb, GG | Right temporal pole | 1a | *BRAF* (c.1799T>A, p.Val600Glu) | Pathogenic | 0.001 | 28 | NM_004333.6 | ENST00000288602.11 | Exome | Y |
| 20 | Austin | FCDIIIb, GG, HS | Left mesial temporal | 1a | *BRAF* (c.1799T>A, p.Val600Glu) | Pathogenic | 0.001 | 18 | NM_004333.6 | ENST00000288602.11 | Exome | Y |
| 21 | Austin | FCDIIa | Right parietal / precuneus | 2b | *NPRL3* (c.275G>A, p.Arg92Gln) | VUS | 0.018 | Germline (Candidate) | NM_001077350.3 | ENST00000611875.5 | Exome | **N** |
| 22 | Austin | FCDI | Right superior and middle frontal gyrus | 4a | - | - | - | - | - | - | Exome, Gene panel | **N** |
| 23 | RCH | FCDI | Right medial frontal/ anterior cingulate | 1a | - | - | - | - | - | - | Exome | **N** |
| 24 | Austin | FCDIa | Left occipital | 4a | - | - | - | - | - | - | Exome | **N** |
| 25 | Austin | FCDIIa | Right post central sulcus, Same region | 4a | - | - | - | - | - | - | Exome | **N** |
| 26 | Westmead | FCDIIa | Left temporal | 3 | - | - | - | - | - | - | Exome, Gene panel | **N** |
| 27 | Austin | FCDIIb | Right ATL and right orbito-frontal, Right anterior cingulate | 1a | - | - | - | - | - | - | Exome | **N** |
| 28 | Austin | FCDIIb | Left inferior frontal sulcus, Same region | 1a | - | - | - | - | - | - | Exome | **N** |

[*]FFPE only.
[†]Previously reported cases: Case 3&4 (Bennett et al., 2022), Case 6 (Bizzotto, S. 2025), Case 8 (Leventer, R.J. 2015)

**Supplementary Table 2.** High-depth Austin Pathology panel sequencing gene list (n = 140)

| MTOR | RAS | | Focal Epilepsy | Cancer | | | | |
|---|---|---|---|---|---|---|---|---|
| *AKT1* | *ALK* | *MAP2K1* | *ATP2A1* | *ACVR2A* | *CTNNB1* | *IDH2* | *NCOA4* | *SDC4* |
| *AKT3* | *BARD1* | *MAP3K3* | *GNA11* | *AIM2* | *DAXX* | *IL6ST* | *NR21* | *SDHA* |
| *DEPDC5* | *BRAF* | *MDM2* | *GNAQ* | *APC* | *DEPDC5* | *KBTBD4* | *NR24* | *SDHB* |
| *MTOR* | *BRCA1* | *MET* | *GRIN2C* | *ARID1A* | *DICER1* | *KDM6A* | *PALB2* | *SDHC* |
| *NPRL2* | *BRCA2* | *MYB* | *KLHL22* | *ASTE1* | *DROSHA* | *KEAP1* | *PBRM1* | *SDHD* |
| *NPRL3* | *BRIP1* | *MYC-N* | *SF3B1* | *ATM* | *EIF1AX* | *KIF5B* | *PDGFRA* | *SLC34A2* |
| *PIK3CA* | *CBL* | *NF1* | *SLC35A2* | *ATR* | *ELOC* | *KIT* | *PMS2* | *SMAD4* |
| *PIK3R1* | *CCND1* | *NF2* | | *ATRX* | *EML4* | *KMT2C* | *POLE* | *SMARCA4* |
| *PTEN* | *CDK4* | *NRAS* | | *BAP1* | *EPCAM* | *KMT2D* | *PPFIA4* | *TAF1B* |
| *RHEB* | *EGFR* | *PRKCA* | | *BAT25* | *EZR* | *MARCKS* | *PRKAR1A* | *TEK* |
| *TSC1* | *ERBB2* | *PTPN11* | | *BAT26* | *FANCL* | *MEN1* | *PRKD1* | *TERT* |
| *TSC2* | *FANCA* | *RAF1* | | *CCDC6* | *FH* | *MITF* | *PTHLH* | *TFEB* |
| | *FGFR1* | *RB1* | | *CD74* | *FOXL2* | *MLH1* | *RAD51A* | *TGFBR2* |
| | *FGFR2* | *RIT1* | | *CDK12* | *GNAS* | *MONO27* | *RAD51B* | |
| | *FGFR3* | *ROS1* | | *CDKN2A* | *H3-3A* | *MRE11* | *RAD51C* | |
| | *HRAS* | *SOS1* | | *CDKN2B* | *H3C2* | *MSH2* | *RAD54L* | |
| | *KRAS* | *STK11* | | *CHEK1* | *HNF1A* | *MSH6* | *RET* | |
| | *LZTR1* | *TP53* | | *CHEK2* | *IDH1* | *NBN* | *RNF43* | |

**Supplementary Table 3.** *NPRL3* (NM_001077350.3): c.393+4_393+7del primer details. Primer set produces two products targeting two *NRPL3* transcripts; 465bp (includes 75bp exon 5), 390bp (excludes 75bp exon 5).

| | Sequence (5’ > 3’) | Template Strand | Length | Tm | GC% |
|---|---|---|---|---|---|
| Forward | ACGGCGATTCCAGGTTTTCA | Plus | 20 | 60.25 | 50.00 |
| Reverse | ACGTGCACAGGCTGTCATAA | Minus | 20 | 59.97 | 50.00 |
| Product Length | 465 | | | | |

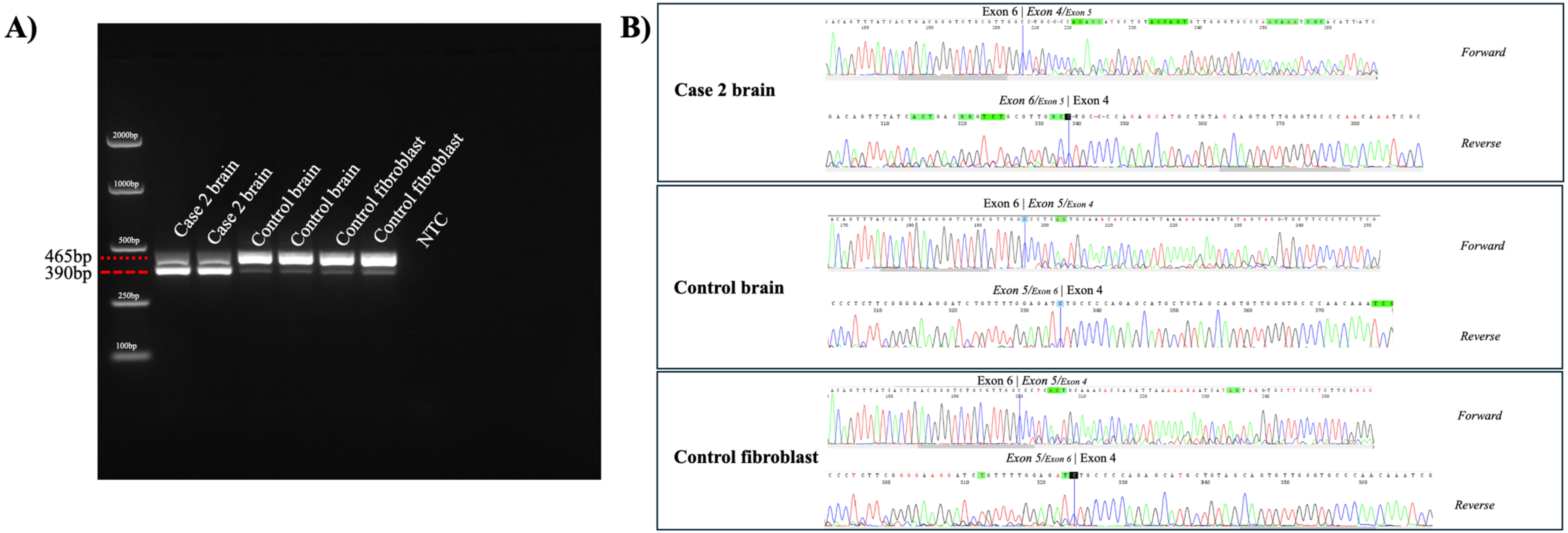


**Supplementary Figure 1.** RT-PCR analysis demonstrates aberrant splicing of *NPRL3* with preferential exclusion of exon 5 in brain-derived RNA from Case 2. A) Gel electrophoresis of RT-PCR products amplified with primers flanking exon 5 (Supplementary Table 3) shows the dominant *NPRL3* transcript excludes exon 5 in the variant carrier, unlike control brain tissue or control fibroblasts. B) Sanger sequencing of RT-PCR products confirms exon 5-containing transcripts are reduced in the variant carrier compared to controls.
